# Does Robotic-Assisted Surgery improve outcomes of total hip arthroplasty compared to manual technique? A protocol for systematic review and meta-analysis

**DOI:** 10.1101/2021.08.05.21261646

**Authors:** Vishal Kumar, Sandeep Patel, Vishnu Baburaj, Rajesh Kumar Rajnish, Sameer Aggarwal

**Author notes:** <u>*Correspondence to:*</u> Dr. Vishnu Baburaj, Senior Resident, Department of Orthopaedics, Postgraduate Institute of Medical Education and Research, Chandigarh, India. 160012. Joint first authors (Vishal Kumar, Sandeep Patel).

## Abstract

**Background:** Robot-assisted total hip arthroplasty (THA) is an emerging technology that claims to position THA components with a very high degree of accuracy. It is unclear if this increase in accuracy leads to improved long-term functional outcomes of the patient.

**Objectives:** This systematic review aims to compare robot-assisted THA to those done using conventional manual techniques, in terms of both short-term radiographic outcomes, as well as long-term clinical outcomes.

**Methods:** This systematic review will be conducted according to the PRISMA guidelines. A literature search will be conducted on the electronic databases of PubMed, Embase, Scopus, and Ovid with a pre-determined search strategy. A manual bibliography search of included studies will also be done. Original articles in English that directly compare robot-assisted THA to manual THA will be included. Data on outcomes will be extracted from included studies and analysis carried out with the help of appropriate software.

## 1. Background

Total hip arthroplasty (THA) is a routinely performed surgery for various diseases of the hip. Its outcomes are linked to the accurate placement of acetabular and femoral components [1]. Complications such as dislocation, impingement, reduced hip movement, accelerated wear of components, aseptic loosening, etc have been associated with improper placement of implants. Several robotic systems have been developed recently to enhance surgical precision during THA, and many surgeons face difficulties in finding merit in the marking claims by various companies [2].

## 2. Need for review

Although there is evidence in literature that robot-assisted techniques lead to improved surgical accuracy, it remains unclear if this would lead to better long-term functional outcomes. This systematic review will shed more light on both the short term as well as long term outcomes of robot-assisted THA, and comparison with conventional manual THA.

## 3. Objective

To compare outcomes of primary THA done with robot-assisted techniques with those done using the conventional manual technique.

## 4. PICO framework for the study

a. Participants : Adult human subjects undergoing primary THA
b. Intervention : Robot-Assisted techniques for THA
c. Control : Manual THA
d. Outcome : Accuracy of component placement, residual limb length discrepancy, surgical duration, complication and revision rates, functional outcomes at final follow-up

## 5. Methods

This systematic review and meta-analysis will be done according to the Preferred Reporting Items for Systematic Reviews and Meta-analysis (PRISMA) guidelines.

### a. Review Protocol

A protocol of the review will be prepared as per the PRISMA-P guidelines.

### b. Eligibility Criteria

Original research on human adult subjects having hip disease undergoing primary total hip arthroplasty (THA) with robotic assistance will be included. The studies should have a comparison group in which primary THA is done using manual techniques. The articles must mention clinical or radiological outcome parameters. Studies in languages other than English, lower evidence studies such as case reports, case series, animal, cadaveric and biomechanical studies will be excluded.

### c. Information Sources and Literature search

Electronic databases of PubMed, Embase, Scopus, and Ovid will be searched using the keywords “Robot* AND (Ortho* OR bone OR hip OR arthroplasty)” for studies in English published from inception to date of search. A bibliography search of included studies will also be carried out for more potentially eligible articles.

### d. Study Selection

Two authors will separately go through the title and abstract of the search results to narrow down studies using the inclusion and exclusion criteria. In case of any doubt, the full text of the study will be obtained and an assessment made after discussion with all the authors.

### e. Data Collection and Data Items

Data from eligible studies will be extracted on excel spreadsheets, which will be cross-checked for accuracy. The following data will be collected:

- Name of first author and publication year
- Study design
- Type of robot used
- Number of participants and their demographic data
- Mean operating time
- Radiographic parameters of components including cup inclination, anteversion, etc.
- Mean limb length discrepancy
- Rate of complications and revision surgeries
- Clinical outcome at final follow-up in terms of functional scoring

### f. Outcome measures

The outcome measures that would be considered for analysis are as follows:

- Accuracy of component positioning
- Mean operating time
- Mean residual limb length discrepancy
- Complication rates
- Rate of revision surgery
- Functional scoring at final follow-up

### g. Data Analysis and Synthesis

Both qualitative and quantitative synthesis will be performed, if adequate data is obtained. Meta-analysis would be conducted to compare the pooled estimate of outcomes between robotic-assisted and manual techniques if reported by more than three studies. RevMan version 5.4 (computer program) will be used for analysis. A fixed or random-effects model will be chosen based on the amount of heterogeneity. 95% confidence intervals will be used, and results would be depicted using forest plots.

### h. Assessment of Risk of Bias

The methodological index for non-randomized studies (MINORS) tool will be used to assess bias in observational studies, and RoB 2.0 tool will be utilized for randomized controlled trials [3, 4].

## Data Availability

All data will be available in the manuscript itself.

